# On the Role of Artificial Intelligence in Medical Imaging of COVID-19

**DOI:** 10.1101/2020.09.02.20187096

**Authors:** Jannis Born, David Beymer, Deepta Rajan, Adam Coy, Vandana V. Mukherjee, Matteo Manica, Prasanth Prasanna, Deddeh Ballah, Michal Guindy, Dorith Shaham, Pallav L. Shah, Emmanouil Karteris, Jan L. Robertus, Maria Gabrani, Michal Rosen-Zvi

**Author notes:** Corresponding authors: **Error! Hyperlink reference not valid.**. **Author’s contributions:** MRZ, DB, JLR, EK and MGA conceived the presented work. MRZ conceived the meta-analysis and supervised this project, and MRZ and MGA set the high-level objectives of this work. JB and MM developed the software to perform paper keyword searches. JB, DB, DR, AC, VM, EK and MGA manually reviewed papers. JB analyzed the results and JB and DR created the figures. All authors contributed toward the interpretation and improvement of the analysis. DB and JB led and distributed the manuscript writing efforts. JB, DB, AC, DR, MGA, EK, DS, PS, MGU and MRZ wrote full, individual sections of the manuscript. All authors reviewed initial versions and contributed significantly to the different sections of the manuscript and approved the submitted version. **Funding Information:** None.

## Abstract

The global COVID-19 pandemic has accelerated the development of numerous digital technologies in medicine from telemedicine to remote monitoring. Concurrently, the pandemic has resulted in huge pressures on healthcare systems. Medical imaging (MI) from chest radiographs to computed tomography and ultrasound of the thorax have played an important role in the diagnosis and management of the coronavirus infection.

We conducted the, to date, largest systematic review of the literature addressing the utility of Artificial Intelligence (AI) in MI for COVID-19 management. Through keyword matching on PubMed and preprint servers, including arXiv, bioRxiv and medRxiv, 463 papers were selected for a meta-analysis, with manual reviews to assess the clinical relevance of AI solutions. Further, we evaluated the maturity of the papers based on five criteria assessing the state of the field: peer-review, patient dataset size and origin, algorithmic complexity, experimental rigor and clinical deployment.

In 2020, we identified 4977 papers on MI in COVID-19, of which 872 mentioned the term AI. 2039 papers of the 4977 were specific to imaging modalities with a majority of 83.8% focusing on CT, while 10% involved CXR and 6.2% used LUS. Meanwhile, the AI literature predominantly analyzed CXR data (49.7%), with 38.7% using CT and 1.5% LUS. Only a small portion of the papers were judged as mature (2.7 %). 71.9% of AI papers centered on disease detection.

This review evidences a disparity between clinicians and the AI community, both in the focus on imaging modalities and performed tasks. Therefore, in order to develop clinically relevant AI solutions, rigorously validated on large-scale patient data, we foresee a need for improved collaboration between the two communities ensuring optimal outcomes and allocation of resources. AI may aid clinicians and radiologists by providing better tools for localization and quantification of disease features and changes thereof, and, with integration of clinical data, may provide better diagnostic performance and prognostic value.

## 1 Introduction

The COVID-19 pandemic has created a desperate need for fast, ubiquitous, accurate, and low-cost tests, and lung imaging is a key complementary tool in the diagnosis and management of COVID-19^1,2^. According to the ACR and the Fleischner Society Consensus Statement, imaging of COVID-19 is indicated in case of worsening respiratory symptoms, and, in a resource-constrained environment, for triage of patients with moderate-to-severe clinical features and a high probability of disease^3,4^. This involves two main tasks. The first is diagnosis, including incidental diagnosis and providing support-evidence in clinical situations in which a false negative RT-PCR test is suspected. The second task is to help evaluate treatment outcomes, disease progression and anticipated prognosis. The field of AI in MI is growing in the context of COVID-19^5,6,7^, and hopes are high that AI can support clinicians and radiologists on these tasks. In this paper, we review the current progress in the development of AI technologies for MI to assist in addressing the COVID-19 pandemic, discuss how AI meets the identified gaps and share observations regarding the maturity and clinical relevancy of these developments.

### 1.1 State of Artificial Intelligence in Radiology

Radiologists play a crucial role in interpreting medical images for the diagnosis and prognosis of disease. Although AI technologies have recently demonstrated performance that matches radiologists’ accuracy in a number of specific tasks, it remains unclear if radiologists who adopt AI-assistance will replace those that do not. As Celi et al. (2019) put it, “*the question is not whether computers can outperform human in specific tasks, but how humanity will embrace and adopt these capabilities into the practice of medicine*”^8^. A steppingstone toward this long-term vision however is the development of AI models that can compete with humans on specific tasks and a pioneer in that progress is the tremendous success in using AI for detection of breast cancer in screening mammography^9, 10, 11, 12^; a success reported by multiple research groups, achieved after 10 years of effort and crowned by OPTIMAM, a database with a total cohort of >150,000 clients^13^.

Similarly, before 2020, significant progress has been made in diagnosing lung conditions using chest X-rays (CXR) and computed tomography (CT), driven by access to publicized annotated datasets. For example, DL-based approaches outperform radiologists in detecting several pulmonary conditions from CXR^14^ and malignancy of lung nodules in low dose CT^15^. Recently, technologies aiming to assist radiologists in such tasks have been made available in the market^16^. However, several key challenges limit the feasibility of adopting these solutions in practice, namely: (i) poor model generalization due to systemic biases, (ii) lack of model interpretability and (iii) non-scalable image annotation processes. Interestingly, similar observations were revealed in the study at hand.

### 1.2 Motivation and Contributions

The recent acceleration of publications intersecting AI and imaging for COVID-19, brings a need for rigorous comparative evaluation of papers to summarize and highlight trends to a broad clinical audience. Previous review papers on COVID-19 either focused on a technical assessment of AI in imaging^6^ or elaborated on the role of imaging^1^. Related systematic reviews were either not devoted specifically to imaging^,17,18^ or used extremely small sample sizes (N=11)^19^. In contrast, this paper attempts to bridge clinical and technical perspectives by providing a comprehensive overview to guide researchers towards working on the most pressing problems in automating lung image analysis for COVID-19.

This is achieved by providing, to date, the largest systematic meta-analysis of AI in MI of COVID-19. Manually analysing 463 publications throughout all of 2020, we attempt to draw a cohesive picture on the current efforts in the field and highlight future challenges, especially related to the cooperation of clinicians and AI experts. While we focus on the lung as the primary organ of SARS-CoV-2 infection, we note the significance of extrapulmonary manifestations^20^.

## 2 Methods

To discover trends from the overwhelming research activities in COVID-19, AI and MI, a systematic review and meta-analysis were performed according to the PRISMA guidelines^21^. Literature, indexed in PubMed and three preprint servers, namely, arXiv, bioRxiv and medRxiv, were queried. The process is illustrated in Figure 1(left) and shows two main streams of queries: a broad one using “AI” AND “COVID-19” AND “Medical Imaging” and a modality-specific one with “AI” AND “COVID-19” AND “Lung” AND (“CT” OR “CXR” OR “US”). Following PRISMA guidelines, we combined the results of both queries across all databases leading to the identification of 463 papers about AI on lung imaging for COVID-19. These papers were included in a manual meta-analysis to review the maturity of the AI technologies and the trends in the rapidly evolving field (for the detailed procedure and a list of synonyms used see appendix Table A1). The publications about AI technology typically tend to report a proof of concept, an illustration of a success in a non-clinical setting, or a report of clinically successful experiments. Additionally, many of the papers identified were not published in peer reviewed journals. To evaluate the maturity of papers, we included five criteria that were assessed rigorously (Figure 1(right)).

**Fig. 1:**
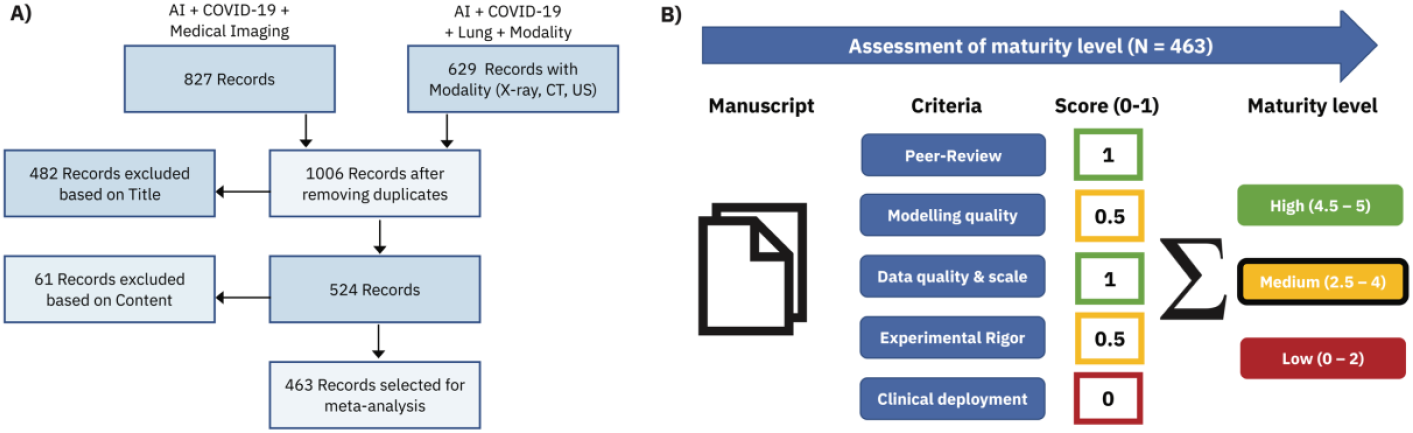
Overview about systematic review and meta-analysis. **A)** PRISMA flowchart illustrating the study selection used in the systematic review. Publication keyword searches on PubMed, arXiv, biorXiv and medRxiv for all of 2020 were performed using two parallel streams. After duplicate matches were removed, titles were screened manually and a selection of 463 relevant manuscripts were chosen for manual review. **B)** Flowchart for quality/maturity assessment of papers. Each manuscript received a score between 0 and 1 for five categories. Based on the total grade, a low, medium or high maturity level was assigned. Details on the scoring system and scores for individual papers can be found in the supplementary material.

1. **Peer review:** Whether or not the paper appeared in a peer-reviewed journal or conference.
2. **Modelling quality:** The complexity and the performance of the developed AI framework.
3. **Data quality/scale:** Number of patients in the data used for training and evaluation. Internal, clinical data is preferred over public datasets and multi-hospital/multimodal data is valued.
4. **Experimental rigor:** Stringency in the evaluation and comparison of the methodology.
5. **Clinical deployment:** The deployment and adoption of the solution in hospitals. Comparison studies of AI and radiologists or deployment of web services were also rewarded.

The peer review score was binary and all other categories were scored ternarily (0, 0.5, 1). Details on the scheme with examples can be found in the supplementary material. The decision function for maturity level (Figure 1 (right)) guarantees that publications that received a “0” in one of the 5 categories cannot get a high maturity score (implying that e.g., preprints are never highly mature). Moreover, we manually inferred the most common tasks addressed in the AI papers, such as detection, segmentation, characterization and outcome prediction, and mapped them into three main clinically relevant categories: diagnosis, severity assessement and prognosis, and one technical task: segmentation. The segmentation papers discuss localization of lung tissue or other disease features without direct applications to any clincially relevant downstream tasks.

For publications that focused on several categories, we consider the primary task only. For example, a number of publications classified as “diagnosis” or “severity assessment” utilized segmentation methods on the fly. Papers that provided a review of ML for MI on COVID-19 and did not introduce original new technoglogy were labeled as “review” papers and excludued from the maturity assessment, leading to 437 reviewed papers. The remaining evaluation criteria per publication were imaging modality, country of authors and country of data source. For each paper, we also recorded the total number of citations indicated on Google Scholar as of 28.2.2021 and converted it to the monthly citation rate. Note that the meta-analysis was blindfolded to the number of citations.

The publication keyword search was performed using our toolbox *paperscraper* that was developed during this project and is open-sourced^*^.

## 3 Results

### 3.1 Progress in AI for Medical Imaging

In recent years, AI solutions have shown to be capable of assisting radiologists and clinicians in detecting diseases, assessing severity, automatically localizing and quantifying disease features or providing an automated assessment of disease prognosis. AI for MI has received extraordinary attention in 2020, as attested by a multitude of interdisciplinary projects attempting to blend AI technologies with knowledge from MI in order to combat COVID-19. A keyword search combining AI and MI revealed 2563 papers in 2019, while 2020 has seen more than twice such papers (5401, cf. Figure 2). Out of these publications, 827 are related to COVID-19, indicating that COVID-19 has accelerated the development of AI in MI.

**Fig. 2:**
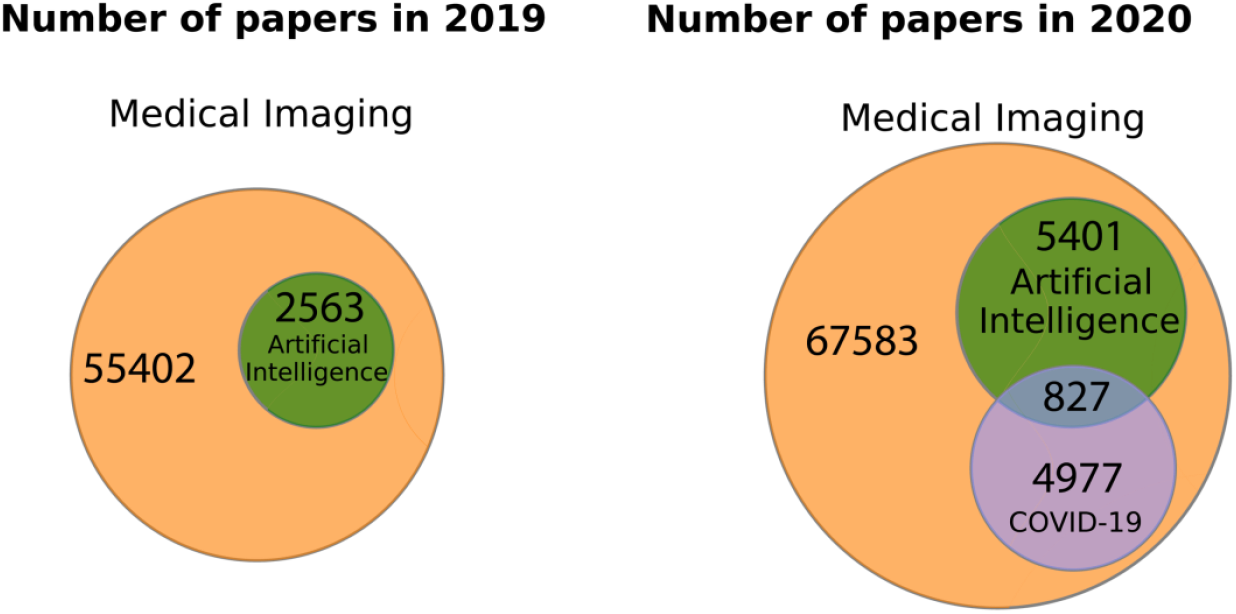
Venn diagrams for AI in MI. Medical imaging received growing attention in 2020, at least partially due to the COVID-19 pandemic. Automatic keyword searches on PubMed and preprint servers revealed that AI has been a majorly growing subfield of MI and that 827 publications in 2020 mentioned the terms MI, AI and COVID-19.

#### 3.1.1 Lung and breast imaging comparison

To enable a perspective on the emergence of AI for MI of COVID-19, we have compiled a comparison on the progress of automatic analysis in breast and lung imaging, as defined in the literature above, from between 2017 and 2020. Figure 3(left) shows a stable growth of papers in AI on both lung and breast imaging over the years 2017-2019. In 2020, the rise of lung-related papers has been accelerated by COVID-19 with a doubling in the first half of 2020 compared to H2 2019 as well as a doubling of 2020 compared to 2019, whereas the trend on AI on mammography imaging remained unaltered compared to previous years.

**Fig. 3:**
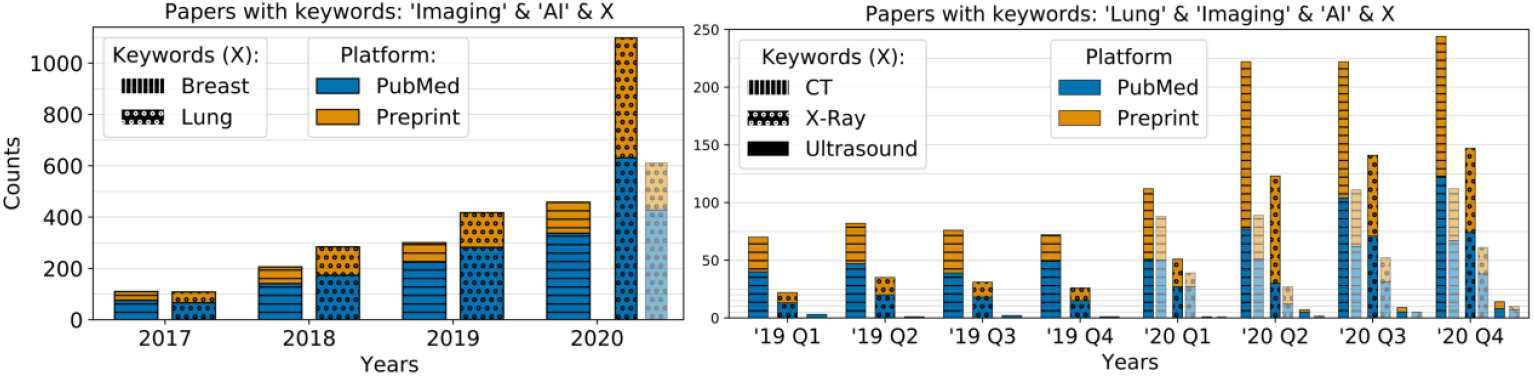
Number of papers per keyword and platform. **Left:** Paper counts using AI on breast or lung imaging. At half-year resolution, the trends persisted; a >100% growth rate for lung was visible in H1 2020 whereas H2 brought about an additional growth of approximately one third (not shown). The lightly shaded bars exclude COVID-19 related papers, which shows the continuity of publications without COVID-19. **Right**: Paper counts comparing the usage of AI on lung imaging modalities. COVID-19 is accompanied by a shift toward more CXR compared to CT papers. For each keyword, multiple synonyms were used (for details see appendix Table A1).

#### 3.1.2 Lung imaging modality comparison

To compare the impact of individual modalities, Figure 3 (right) shows that 2019 witnessed a stable trend of ∼100-120 papers per quarter on AI whereas with the COVID-19 outbreak in 2020, numbers soared to 164, 352, 372 and 405 papers for Q1-Q4, 2020 respectively. This rise was spontaneously evoked by COVID-19, as excluding papers mentioning COVID-19 would have resulted in a continuation of the stable trend (see lightly shaded bars) of a hypothetical ∼120-160 publications. Notably, the relative contributions of the modalities changed toward CXR from 2019 to 2020 (shares of 71% vs 63% for CT, 27% to 35% for CXR and 2% for US respectively). Moreover, for non-COVID-19 papers, the ratio between preprints and PubMed indexed papers for AI in breast and chest is 29% and 37% from 2017-2019, respectively; for COVID-19 related papers, this ratio rose to 58%.

### 3.2 Broad Insights from Meta-analysis

By focusing on CT, CXR and US, we quantified the publication efforts of AI for MI of COVID-19 and identified 463 papers which were included in a manual meta-analysis to review the maturity of the AI technologies and the trends in the rapidly evolving field. The full spreadsheet with individual scores for each publication is available in the supplementary material.

#### 3.2.1 Disparity between Clinical and AI Communities

Of the 4977 papers about MI and COVID-19 (see Figure 2 (right)), 2496 are specific to modalities as shown in Figure 4 (left), indicating a dominance of CT in clinical papers (84%), followed by CXR (10%) and LUS (6%). By using publication counts as an indirect indicator on scientific response, we observe a mismatch in the focus of the AI community in comparison to the clinical community as illustrated by the distribution of papers per modality in Figure 4 (right) that shows a clear dominance of CXR (50%) across AI papers.

**Fig. 4:**
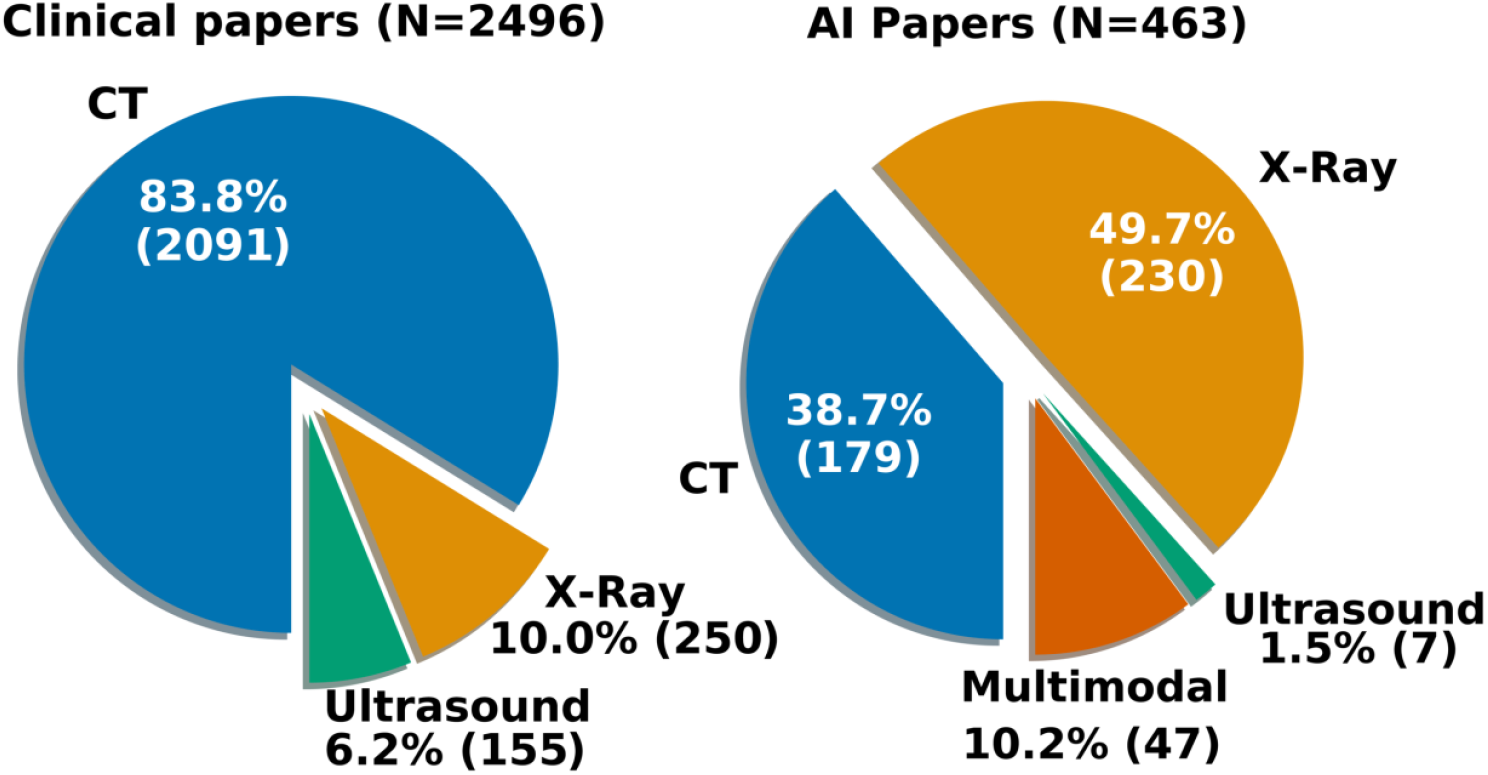
Imaging modality comparison during the COVID-19 pandemic. CT takes the lion’s share of clinical papers about lung imaging of COVID-19 (left). The AI community (right) instead published disproportionally more papers on CXR compared to clinicians, whereas CT and also Ultrasound are underrepresented. Multimodal papers used more than one imaging modality.

In addition, the vast majority (72%) of papers focused on diagnosis of COVID-19 over tasks like severity and prognosis (Figure 5 (left)). This trend is in contrast to the ACR guidelines appraising imaging as an inconclusive test for COVID-19 detection due to uncertainties in accuracy and risk of cross-contamination. Revealing was the unanimous use of CXR data (50%, see Figure 4 (right)) that was commonly utilized without any further clinical or radiomic features. The tendency for diagnosis was especially prominent for CXR versus CT where 87% and 58% diagnosis papers were found respectively (cf. the sunburst plot showing task and maturity as distributed by modality in the supplementary material, Fig. A1). While 6% of papers (27 of all 437 non-review papers) exploited multimodal imaging data towards building their AI models, studies on multimodal imaging data of the same patient cohort are lacking with few exceptions. In one example manual disease airspace segmentation from CT was used as ground truth for volumetric quantification from CXR^22^. Another study demonstrated the diagnostic accuracy (ROC-AUC) of AI on CT to be clearly superior to CXR^29^.

**Fig. 5:**
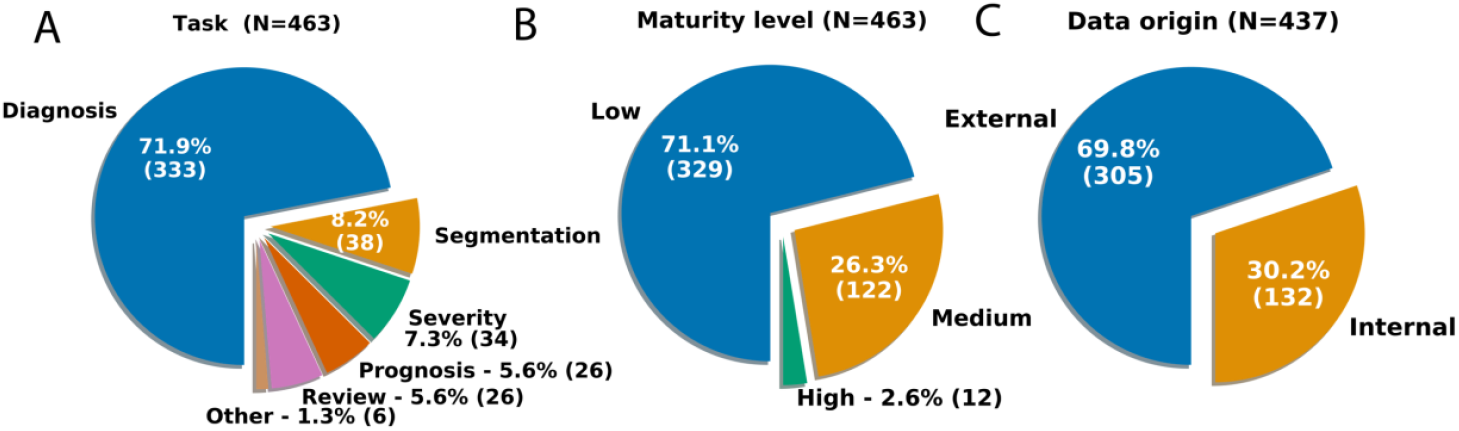
Distribution of manually reviewed papers on AI and medical imaging during the COVID-19 pandemic. Relative proportions for primary performed task (left), quality (middle) and data origin (right) are given. N is smaller for B) and C) since review papers were excluded from that analysis.

#### 3.2.2 Most AI Solutions for COVID-19 have Low Maturity

The maturity of the papers was assessed following the scheme in Figure 1 (right) by co-authors who have developed or worked with DL algorithms (see Figure 5 (middle)). Almost 70% of papers were assigned a low maturity level and only 12 (2.7%) highly mature studies were identified. A detailed spreadsheet with the evaluations of each paper is included in the supplementary material.

CT papers had a higher maturity score than CXR papers (2.1 +-1.3 vs. 1.3 +-1.1, *p*<1e-11, MWU) and 57% of CT versus 43% of CXR papers were peer-reviewed. As the pandemic continues the preprint ratio is declining steadily (from 69% in Q1 to 45% in Q4) but not (yet) significantly (r=-0.93, *p*=0.07). The maturity score also heavily varies across performed task and was significantly higher for COVID-19 severity assessment and prognosis (2.4 and 2.5) compared to diagnosis/detection (1.5) and segmentation (1.6) as assessed by post-hoc Tukey’s HSD multiple comparison tests (see Figure 6).

**Fig. 6:**
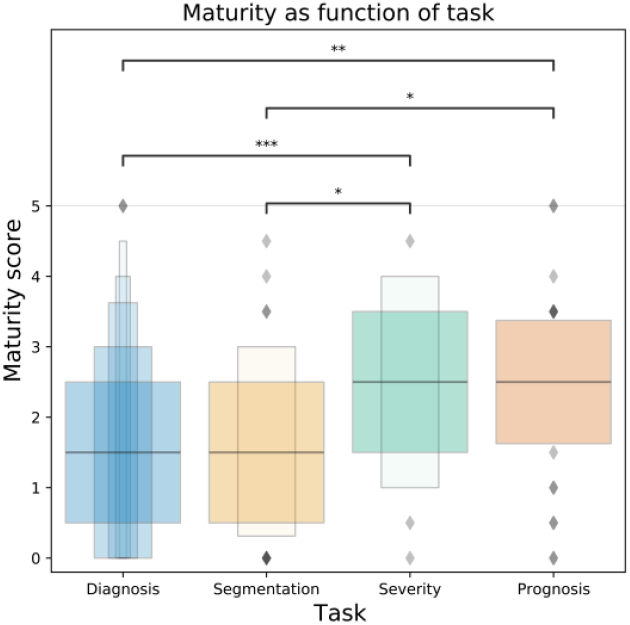
Maturity score as function of task (N=437). Publications focusing on COVID-19 diagnosis/detection or pure segmentation achieved a significantly lower maturity score than publications addressing /severity assessment/monitoring or prognostic tasks (stars indicate significance levels 0.05, 0.01 and 0.001, respectively).

Posteriori, we observed that the monthly citation rate was significantly greater for 1) high compared to medium maturity papers (6.9 vs. 2.3, *p*<0.01, U) and 2) medium compared to low maturity (2.3 vs. 1.9, *p*<0.05, U). The continuous maturity score was found to be significantly correlated (r=0.12, p<0.05) with the monthly citation rate. Interestingly however, a major factor accounting for a high citation rate is not the maturity but the months elapsed since publication (r=0.35, *p*<1e-14). This suggests that absolute citations and relative citation rates are insufficient quality measures and we instead observe a tendency towards continuous citation of publications that appeared early in the pandemic (irrespective of their quality).

#### 3.2.3 Overuse of Small Incomprehensive Public Datasets

We observed that only 30% of papers used proprietary or clinical data (Figure 5 (right)), while almost 70% analysed publicly available databases. Such databases exist for CT^23^, CXR^24^ and LUS^25^ and are usually assembled by AI researchers, contain data fetched from publications and are comprised of no more than a few hundred patients from heterogenous sources/devices without detailed patient information. Accordingly, the geographical diversity of data sources was not extremely high (26 countries), and by a wide margin the three most important data donators were countries hit early from the pandemic, namely, China (48%) and, to a lesser extent, USA (12%) and Italy (11%). Interestingly, a global collaborative spirit towards combatting COVID-19 was revealed as first-authors from 53 countries and 6 continents contributed to the research – with the most active countries being China (21%), USA (13%) and India (11%).

### 3.3 Uncovering Trends in AI Solutions from the Mature Papers

12 (2.7%) of the assessed papers were assigned high maturity ^26,27,28,29,30,31,32,33,34,35,36,37^. The list of papers together with details about their task, key finding, implementation and results appear in Table 1 and are further discussed in this section.

**Table 1.** Details on the 12 best papers found in our systematic meta-review of 463 papers (maturity score of high).

| Paper Title | Primary Task & Modality | Key Findings | Limitations | Patients (train/val/test) | Number of Data Sites | Labels | Architecture, Dimensionality | Pretraining | Metrics | Results | Reproducibility (code/data open source) |
| --- | --- | --- | --- | --- | --- | --- | --- | --- | --- | --- | --- |
| Artificial intelligence-enabled rapid diagnosis of patients with COVID-19. [27] | Diagnosis – CT | System identified 68% of RT-PCR positive patients with normal CT (asymptomatic). Clinical information is important for diagnosis and model is equally sensitive than a senior radiologist. | Small data size, mild cases have few abnormal findings on chest CT, severity of pathological findings variable in CT. | 534<br>92<br>279 | 18 | RT-PCR tests | Inception-ResNet-v2 (pre-trained ImageNet), 3-layer MLP, 2D | Transfer learning (pulmonary tuberculosis model) | AUROC, Sensitivity, Specificity | 0.92 AUC, 84.3% Sens. 82.8% spec | Code - Yes, Data - No |
| Artificial Intelligence Augmentation of Radiologist Performance in Distinguishing COVID-19 from Pneumonia of Other Origin at Chest CT [33] | Diagnosis – CT | AI assistance improved radiologists' performance in diagnosing COVID-19. AI alone outperformed radiologists on sensitivity and specificity. | Bias in radiologist-annotation, heterogenous data, bias in location of covid (China) vs. non-covid pneumonia patients (US) | 830<br>237<br>119 | 13 | RT-PCR tests, Slice-level by radiologist | EfficientNet-B4, 2D | Transfer learning (ImageNet) | AUROC, Sensitivity, Specificity, Accuracy, AUPRC | 0.95 AUC, 95% Sens, 96% Spec, 96% Acc, 0.9 AUPRC | Code - Yes, Data - No |
| Automated Assessment of CO-RADS and Chest CT Severity Scores in Patients with Suspected COVID-19 Using Artificial Intelligence. [34] | Diagnosis – CT | A freely accessible algorithm that assigns CO-RADS and CT severity scores to non-contrast CT scans of patients suspected of COVID-19 with high diagnostic performance. | Only one data center. High COVID prevalence, low prevalence for other diseases. | 476<br>105 | 1 | RT-PCR, radiology report | Lobe segmentation 3D UNet, Co-rads scoring 3D Inception Net | Transfer learning (ImageNet and kinetics) | AUC, Sensitivity, Specificity | Internal: 0.95 AUC<br>External: 0.88 AUC | Code - Yes, Data - No |
| Diagnosis of covid-19 pneumonia using chest radiography: Value of artificial intelligence [35] | Diagnosis – X-Ray | AI surpassed senior radiologists in COVID-19 differential diagnosis. | High COVID prevalence. Human ROC-AUC were averaged from 3 readers. | 5208<br>2193 | 5 hospitals, 30 clinics | RT-PCR NLP on radiology report | CV19-Net | 3 Stage transfer learning (ImageNet) | AUC, Sensitivity, Specificity | 0.92 AUC, 88.0% Sens. 79.0% spec | Code - Yes, Data - No |
| Development and evaluation of an artificial intelligence system for COVID-19 diagnosis. [29] | Diagnosis – Multimodal | Paired cohort of CXR/CT data: CT is superior to CXR for diagnosis by wide margin. AI system outperforms all radiologists in 4-class classification. | More data on more pneumonias subtypes needed. No clinical information used (could enable severity assessment) | 2688<br>2688<br>3649 | 7 | - | lung seg 2D Unet, slice diagnosis 2D ResNet152, | Transfer learning (pre-trained ImageNet) | AUC, Sensitivity, Specificity | AUC 0.978 | Code - Yes, Data - No |
| AI-assisted CT imaging analysis for COVID-19 screening: Building and deploying a medical AI system. [31] | Diagnosis – CT | System was deployed in 4 weeks in 16 hospitals AI outperformed radiologists in sensitivity by wide margin. | Model fails when multiple lesions, metal or motion artifacts are present. System depends on fully annotated CT data | 1136 | 5 | NAT, 6 annotators (lesions, lung) | 3D UNet++, ResNet50, | Full Training | Sensitivity, Specificity | Sens 97.4%, Spec 92.2% | Code - No, Data - No |
| Automated assessment and tracking of COVID-19 pulmonary disease severity on chest radiographs using convolutional siamese neural networks [32] | Severity – X-Ray | Continuous severity score used for longitudinal evaluation and risk stratification (admission CXR score predicts intubation and death, AUC=0.8). Follow-up CXR score by AI is concordant with radiologist (r=0.74) | Patients only from urban areas in US. No generalization to PA radiographs. | 160,000<br>267 (images) | 2 | RT-PCR tests, 2-5 annotators, mRALE | Siamese DenseNet-121 | DenseNet-121, (ImageNet, fine-tuned on CheXpert) | PXS score, Pearson, AUC | r=0.86, AUC = 0.8 | Code - Yes, Data - Partial (covid CXR not released) |
| Development and clinical implementation of tailored image analysis tools for COVID-19 in the midst of the pandemic [36] | Severity<br>–<br>CT | Developed algorithms for quantification of pulmonary opacity in 10 days. Human-level performance with <200 CT scans. Model integrated into clinical workflow. | Data: no careful acquisition, not complete, consecutively acquired or fully random sample. Empirical HU-thresholds for quantification. | 146<br>66 | 1 | RT-PCR, 3 radiologist annotators | 3D Unet | Full Training | Dice coefficient, Hausdoff distance | dice=0.97 | Code - Yes, Data - No |
| Clinically Applicable AI System for Accurate Diagnosis, Quantitative Measurements, and Prognosis of COVID-19 Pneumonia Using Computed Tomography [26] | Prognosis<br>–<br>CT | AI with diagnostic performance comparable to senior radiologist. AI lifts junior radiologists to senior level. AI predicts drug efficacy & clinical prognosis. Identifies biomarkers for NCP lesion. Data available. |  | 3777 | 4 | Pixel-level annotation (5 radiol.) | Lung-lesion seg DeepLabV3, diagnosis analysis 3D ResNet-18, Gradient boosting decision tree | Full Training | Dice coefficient, AUC, accuracy, sensitivity, specificity | 0.9797 auc, 92.49% acc, 94.93% sens, 91.13% spec, | Code - Yes, Data - Yes |
| Relational modelling for robust and efficient pulmonary lobe segmentation in CT scans [30] | Segmentation<br>–<br>CT | Leverages structured relationships with non-local module. Can enlarge receptive field of convolution features. Robustly segments COVID-19 infections. | Errors on border of segmentations. Gross pathological changes not represented in data. | 4370<br>1100 | 2<br>(pretraining: 21 centers) | Radiology report | RTSU-Net (2-stage 3D Unet) | Pre-training on COPDGene | Intersection Over Union, Average Asymmetric Surface Distance | IOU 0.953, AASD 0.541 | Code - Yes, Data – No/partial |
| Dual-branch combination network (DCN): Towards accurate diagnosis and lesion segmentation of COVID-19 using CT images. [37] | Diagnosis<br>–<br>CT | DCN for combined segmentation and classification. Lesion attention (LA) module improves sensitivity to CT images with small lesions and facilitates early screening. Interpretability: LA provides meaningful attention maps, | Diagnosis depends on accuracy of segmentation module. No slice-level annotation. | 1202 | 10 | RT-PCR, pixel level annotation by 6 radiologists | UNet, ResNet-50 | Full Training | accuracy, dice, sensitivity, specificity, AUC, average accuracy | acc 92.87%, dice 99.11%, sens 92.86%, spec 92.91%, 0.977 AUC, 92.89% Avg Acc | Code - No, Data - No |
| AI-driven quantification, staging and outcome prediction of COVID-19 pneumonia. [28] | Prognosis<br>–<br>CT | 2D/3D COVID-19 quantification, roughly on par with radiologists. Facilitates prognosis/staging which outperforms radiologists. Rich set of model ensembles, uses clinical features. | Test dataset partly split by centers. | 693<br>(321k slices)<br>513 for test | 8 | RT-PCR | AtlasNet, 2D | Full Training | Dice coefficient, correlation, accuracy | Dice 0.7, balanced accuracy 0.7 | Code – No Data – Yes (without images) |

We summarize the trends observed in the identified list of mature papers with a deeper focus on aspects such as: (i) choice of AI model architecture, (ii) diversity in data sources, (iii) choice of evaluation metrics, (iv) model generalization and (v) reproducibility. Further, we highlight common limitations reported in these papers.

i. **AI Modelling**: Most of the presented AI solutions have high complexity comprising of multiple modelling stages with at least 2 models and at most an ensemble of 20 models^35^ being trained. Solutions for segmentation tasks tend to model 3D data, while classification tasks used 2D data. Almost all of the solutions used transfer learning with pre-training on ImageNet or other open-source clinical datasets (e.g., CheXpert, COPDGene). Popular neural network architectures used included UNet, ResNet, DenseNet and InceptionNet.
ii. **Data Sources**: The majority of mature publications utilized data obtained from multiple hospitals containing about 500 to 5000 patients’ imaging data. The datasets were typically labelled using manual annotations from radiologists, RT-PCR tests and results from radiology reports. Note that only three studies utilized clinical metadata in addition to images to develop their AI system ^26,27^,28.
iii. **Evaluation Metrics**: The publications addressing diagnosis tasks commonly used metrics such as accuracy, AUC, sensitivity, and specificity to evaluate the model performance, while using dice and intersection over union scores to quantify performance on segmentation tasks. The Pearson correlation coefficient was routinely used to compare model and human reader performances and understand the influence of learned features on the overall system performance.
iv. **Experimental Rigor and Model Generalization**: We observed that while most publications reported confidence intervals and performed statistical tests, they evaluated their algorithm typically only on a single random split of the dataset. Most mature publications reported model performance on external test datasets, as well as presented heatmaps to illustrate regions of image the model focused on. However, few conducted cross-validation and ablation studies to understand the generalization capabilities of their models. Further, a couple of solutions were deployed in clinical practice^31,36^ and another one of them was also thoroughly tested in multiple countries^26^.
v. **Reproducibility**: All of the mature publications used a human-in-the-loop (about 1 to 8 experienced radiologists) to compare and evaluate their proposed AI solutions, thus making such an evaluation scheme a standard practice. Moreover, a majority of the studies released the code for their algorithm publicly, while the data usually remained proprietary, but was at least partly released in four mature papers ^26,28,30,32^.
vi. **Limitations**: All publications acknowledge limitations in their studies owing to inherent biases that are modelled into in the datasets through limited size, lack of diversity, and imbalance in disease conditions. In many situations, the datasets represented population of patients with higher prevalence of COVID-19 at the time of imaging which does not reflect true disease prevalence. Further, the models were deemed sensitive to motion artifacts, and other subtypes of lesions or comorbidities which cause data distribution shifts. Most studies also utilized datasets from limited geographical locations thereby restricting generalization performance of the models in other geographies.

### 3.4 Task-specific Review of Publications

In this section, we discuss the four categories of tasks addressed by the 463 papers chosen for meta-analysis, namely: diagnosis, severity, prognosis and segmentation. We also highlight key results from the 12 mature publications and provide an overview of the findings specific to COVID-19.

#### 3.4.1 Diagnosis

We find that 72% of the papers centered on COVID-19 diagnosis with 8 out of the 12 mature papers (75%) also addressing this task. As the most prominent COVID-19 test relies on the identification of viral RNA using RT-PCR^38^, imaging is not routinely performed/recommended for diagnosis and given its reliance on pulmonary pathologies, it is especially inappropriate for detection of early or asymptomatic infections^39^. However, compared to nucleic acid tests, CTs may be more sensitive at a single time point for the diagnosis of COVID-19^40^. A key diagnostic challenge is the non-specificity of COVID-19 patterns and their differentiation from non-COVID-19 viral pneumonia ^41^. Here, non-imaging assessments like anamnesis can contribute to the diagnosis. Secondly, asymptomatic patients with unaffected lungs are notoriously challenging to be detected. In both cases, however the lack of visibly distinguishing features for COVID-19 might not directly imply a limited ability of DL-based approaches, which might still be able to automatically identify (segment) distinguishing features, given the appropriate data for training^42^. As it has been demonstrated, if DL approaches combine CT and clinical features, the performance of radiologists in the detection of symptomatic COVID-19 patients can be matched^27^ (or surpassed^33^), and even asymptomatic patients with normal CT scans can be identified in 68% of the cases^27^.

Moreover, multiple studies validated that radiologists’ performance improves upon consultation of AI: Junior radiologists along with AI can perform as well as mid-senior radiologists^26^ and radiologists’ sensitivity and specificity can improve by nearly 10% through AI^43^. In another study, AI recovered full-dose CT from ultra-low-dose CTs with a satisfying acceptance score of 4.4 out of 5 by radiologists (compared to 4.7 and 2.8 for full- and ultra-low-dose respectively) and thus helped to reduce the CT radiation dose by up to 89% while still facilitating downstream diagnosis^44^. One highly mature diagnostic study using CXR included almost 6000 scans from >2000 COVID-19 patients and their DL model exceeded the diagnostic performance of thoracic radiologists as found by significantly higher area under the curve of 0.94 (vs. 0.85) and sensitivities when matching specificity to radiologists’ performance^35^.

#### 3.4.2 Severity Assessment

Imaging findings of COVID-19 patients correlate with disease severity^45^ and CT scanning can assess the severity of COVID-19 and help monitor disease transformation among different clinical conditions^46^. A retrospective comparison of imaging findings on chest CTs with disease severity revealed an increased occurrence of consolidation, linear opacities, crazy-paving pattern and bronchial wall thickening in severe patients at a higher frequency than in non-severe COVID-19 patients. The CT findings correlated with several worse symptoms, including a respiratory rate greater than 30 breaths per minute, and oxygen saturation of 93% or less in a resting state among other phenotypes^47^. In clinical practice, often progress assessments as well as patient management is performed based on CXR and not chest CT. AI that provides assessment of severity could be useful if it was quantifiable and accurate but only one publication was found mature in performing this task^36^. The authors developed a clinically useful AI tool consisting of a U-Net backbone for lung segmentation and quantification of pulmonary opacity within 10 days and achieved human-level performance when training on less than 200 CT scans^36^. Another work utilized a dataset of multiple CT scans per patients and introduced a “CT scan simulator” that modelled the temporal evolution of the CT through disease progression and was evaluated on multi-national and multi-machine data^48^. Their work proposed to decompose the task of CT segmentation from one 3D into three 2D problems, thus achieving remarkable performance. Notably, despite the overall overhead of CXR compared to CT in the analysed publications, only 3% (n=6) of the CXR publications in the meta-analysis focused on severity assessment (cf. 14% for CT). One of them trained DL models on lung segmentation and opacity detection of 48 COVID-19 patients and achieved an agreement measure (Cohen’s kappa) of 0.51 for alveolar opacities and 0.71 for interstitial opacities^49^. In one publication with multimodal imaging data for one patient cohort, manual airspace disease segmentation of CTs in 86 COVID-19 patients was used as ground truth to train a super-resolution CNN on volumetric quantification from CXR^22^. The obtained correlation percentage of opacity (PO) volume (CT) and PO area (CXR) was around 0.8 for both AI and averaged human experts. A recent study on LUS first inferred a patient-level representation from the region-level LUS videos using attention-based multiple-instance learning and then performed semi-supervised contrastive learning to integrate imaging with clinical data^50^. The method achieved 75% and 88% accuracy in a 4-level/2-level patient severity assessment respectively and even identified infected regions in LUS (B-lines) en passant.

#### 3.4.3 Prognosis

Very few of the papers (26 i.e., 6%) focused on prognostic assessments of COVID-19 such as treatment outcome prediction, risk assessment (e.g., requirement for ICU admission or mechanical ventilation) or time elapsed to negative PCR. However, two of them were assessed as mature^26,28^ and the average maturity score was the highest for this task (cf. Figure 6). However, in contrast to diagnosis, these tasks are clinically more relevant as they cannot be performed routinely and reliably with standard care. While this can be attributed to an overall gap in knowledge of the long-term effects of COVID-19 and a lack of historical data to enable training on large scale prognosis data, it is constructive towards the alignment of future research in the field. On the other side, in the past few months the hyper-inflammatory response induced by COVID-19 has been identified as a major cause of disease severity and death ^51^. Thus, studies have focused on the identification of predictive biomarkers of pathogenic inflammation. Lung imaging is not expected to reflect these biomarkers’ expression, leading to limited prognosis accuracy based on imaging. One study assessed as highly mature, seamlessly integrated a diagnostic module (based on a CT lung-lesion segmentation) with a prognostic module that combined clinical metadata and quantification of lung-lesion features^26^. The system demonstrated diagnostic performance comparable to a senior radiologist and the prognostic module predicted progression to critical illness and could evaluate drug treatment efficacy by three drugs. Notably, the multi-center dataset of 3,777 patients as well as the source code is available to the public to support the development of a better system and to validate their study.

#### 3.4.4 Segmentation

The main abnormalities observed in common and severe COVID-19 cases are ground glass opacities (GGOs) and patchy consolidation surrounded by GGOs. COVID-19 pneumonia manifests with chest CT imaging abnormalities, even in asymptomatic patients, with rapid evolution from focal unilateral to diffuse bilateral GGOs that progress to or co-exist with consolidations within 1–3 weeks^52^. The visual features of GGOs and consolidation lend themselves to image analysis by DL networks, and with 27 publications (8%) segmentation became the second-most performed task after diagnosis. In our analysis, many of the papers performed segmentation to enable other clinical tasks as discussed above, but one mature study focused on providing a pulmonary lobe segmentation with relational modelling^30^. Using topological modelling techniques that explore structural relationships between vessels, airways and the pleural wall and break up with the common strategy of utilizing fully local modules such as convolutions, they achieved human-level performance. In most cases (82%), segmentation publications utilized external data sources with little or no clinical collaboration. Some segmentation-based models output pixelwise-labelled tissue maps of GGO or consolidation regions, providing quantitative localization of findings and identification of disease features, which can be especially informative in clinical tasks such as grading disease severity or tracking progression over time. Chaganti et al. achieved this by segmenting anatomical landmarks with reinforcement learning and computing percentage of opacity and lung severity score as complementary severity measures^53^.

In an exhaustive empirical evaluation of DL models on a clinical dataset of almost 100 COVID-19 patients, distinguishing lesion types was found more difficult than lung segmentation or binary lesion segmentation while model ensembles demonstrated best performance^54^. The manual delineation from radiologists, valuable for segmentation tasks, inherently introduces some inter-rater variability which underlines the need for segmentation techniques that can deal with uncertainty in annotations^55^.

## 4 Discussion

In summary, the number of papers on AI in MI for COVID-19 has grown exponentially in 2020 and the quality of the manuscripts varies significantly. In our manual review, only 12 (2.7%) highly mature studies were identified. A key characteristic that underpins highly mature studies is an interdisciplinary and often multi-national collaboration of medical professionals and computer vision researchers.

### 4.1 Challenges and possible solutions

Given the observed disparities between the AI and medical communities, we discuss several challenges that are currently encountered in such interdisciplinary collaborations and provide potential approaches to remedy the same.

### 4.2 Choosing the right task for AI models

The AI literature primarily addresses diagnostic tasks as opposed to other tasks with higher clinical relevance, such as monitoring/severity estimation (which tracks with clinical outcomes) and management tasks such as ventilation equipment and bed allocation. Currently, even the best AI solutions have minimal performance gains on well-defined tasks (such as diagnosis) and are thus unlikely to be adopted clinically^8^.

Conclusions from our meta-analysis are that (1) the choice of task is critically driven by the availability of annotated data and (2) the speed of execution in AI propels blind response to increase short-term rewards instead of finding solutions to high-priority problems. This partly explains the over-attention to diagnostic tasks. Moreover, classification is the canonical ML formulation and while regression techniques can estimate non-binary severity scores, they are are less frequently used. Severity estimation can be reduced to summing a classification problem on the pixel level, but this requires very expensive pixelwise annotated training data. Another common misalignment between communities is the disparate objective functions in diagnostic classification of COVID-19 from imaging data. Irrespective of the availability of direct tests for SARS-CoV-2, radiologists around the globe are steered by the objective to avoid false negatives; their decisions are less factious and dichotomous and more granular than a categorical classification of a ML model. On the other hand, the utility of an AI model, trained on a ground truth assigned by radiologists’ interpretation is limited and mostly restricted towards saving time and resources than getting better decisions.

To remedy and develop better clinical prediction models the seven steps for development and four steps for validation proposed by Steyerberg et al. (2014)^56^ should be followed and complemented by an increased motivation among AI experts to focus on the right questions and leverage suitable, and radiologists-friendly, inductive biases like soft labelling^57^. Since AI techniques are data-driven, the best way to steer AI practice towards more COVID-19 clinical relevance is to collect CT data with annotations for severity, as well as demographics data and outcomes data. Recent collaborative, multi-institution data collection efforts such as the NHS NCCID and RSNA’s RICORD data sets precisely have CT data combined with outcomes and severity, and they are sure to lead to AI approaches with more clinical impact. AI challenge competitions are a related route for channelling AI towards CT and severity estimation. MICCAI’s COVID-19 lung CT lesion segmentation challenge collected a CT data set with detailed, radiologist-labelled lesions on the pixel level. AI-based lesion segmentation can then estimate severity by counting lesion voxels. In general, the hope is that this can be applied to longitudinal studies to track COVID-19 progression, and eventually be combined with demographics and hospitalization data. Last, two other promising and clinically relevant endeavours are (1) usage of DL for generating standardized assessment of pulmonary involvement of COVID-19 by leveraging the newly introduced COVID-19 Reporting and Data System (CO-RADS)^34^ and (2) using DL to help evaluate treatment outcomes, e.g., by assessing changes in lesion size and volume changes^26^.

#### 4.1.2 Transparency and reproducibility

While most authors of highly mature studies released their code (indeed three papers did not release code ^28,31,37^) only a third of them released at least part of their data. This raises concerns about reproducibility and transparency of their studies, as recently argued against a *Nature* study on breast cancer screening^11^ in a “*matters arising”*^58^. Similarly, COVID-19 mortality prediction study^59^ was found to be irreproducible by three independent research groups from different countries^60,61,62^. Given the global, unprecedented public health challenge caused by COVID-19, we strongly encourage medical researchers to follow the trends toward **open-source** development in the field of machine learning (which has been proclaimed by various luminaries fourteen years ago^63^ and successfully implemented in important venues). We encourage to expedite a transformation toward a common practice of validating the proposed methodology and results by publishing both code and, whenever possible, anonymized medical data; especially in academic, non-commercial settings. To help foster this transformation, conference organizers and journal editors should encourage the open sharing of code and anonymized data in their call for papers and add this as criterion to the review procedure. For example, NeurIPS and ICML, premier machine learning conferences, expect that submissions include code and anonymized data and take this into account during the decision-making process. Similarly, the imaging conferences CVPR and MICCAI both strongly encourage the inclusion of code and data. Better guidelines from official sources such as the government are needed especially since data sharing regulations are less stringent during a pandemic and medical facilities are often not aware of the numerous advantages of data sharing. Privacy-preserving data science techniques have advanced^64^ and should help to build more trust toward data sharing.

Federated learning (FL) is an emerging realm of ML concerned with distributed, decentralized training that stores privacy-sensitive data only locally (for details see ^65,66,67^). It allows multiple parties to collaboratively train the same model without data sharing and could thus become key to foster collaborations between clinical and AI communities and overcome privacy concerns. Our meta-analysis included three preprints exploring FL using CT^68^ or CXR^69^ data. A recent FL study on EHR from 5 hospitals was found to improve COVID-19 mortality prediction^70^. These efforts will hopefully increase reproducibility and make comparative studies more feasible, which will help the research community focus on the highest performing methods.

#### 4.1.3 Imaging modality rivalry

An ideal imaging modality should be safe, ubiquitous, accurate, fast, and preferably provide high-quality reproducible results via portable devices. The three different imaging modalities, addressed in this study, differ in their clinical use, their availability, portability, safety and reproducibility and none of them is ideal for addressing all aspects of the pandemic (for a comparison see Table 2). Herein, we have unravelled a mismatch in the number of publications per modality between clinical and AI communities – the AI literature has focused mostly on CXR whereas CT and LUS have received comparably little attention (cf. Figure 4). CT is deemed the gold-standard, dominates in clinical publications, is more sensitive than CXR for detecting diseases of the chest, but is restricted to modern medical facilities ^71^. CXR is notoriously less sensitive than CT ^72^, yet it is the most abundantly used modality across the globe when managing COVID-19 patients. While CXR can underestimate disease, CT can narrow down a differential diagnosis that appears broad on CXR. For AI, large data sets are needed for ML approaches, and there are much larger data sets for CXR than for CT.

**Table 2.** Differences between the imaging modalities.

|  | CT | CXR | LUS |
| --- | --- | --- | --- |
| **Benefit** | High sensitivity, high specificity | Fast, broadly available | Mobile, radiation-free, broadly available |
| **Drawback** | Patient transportation<br>Low availability<br>Radiation dose<br>Increased workload for disinfection | Low sensitivity<br>Non-specific<br>Large volume of radiographs leads to increased workload. | User dependent<br>Non-specific<br>Long acquisition time<br>Requires patient interaction |
| **Clinical Role** | Diagnose additional complications<br>Rule out additional etiologies of symptoms (effusions, bacterial pneumonia) | Initial diagnosis<br>Monitoring clinical progression<br>Detection of complications | Triage<br>Point-of-care monitoring for specific tasks |

As the use of imaging is less regulated compared to PCR/antigen testing, an official recognition of all imaging modalities by leading institutions and stakeholders is needed. In conjunction with clear guidelines for clinicians on when to use which modality, trust into imaging can be increased and workflows can be streamlined. For example, the practical advantages of LUS include non-invasiveness and portability and its consequent role in triage^73^. However, LUS is operator dependent and requires close patient contact for a relatively longer time^74^. It was described as a preferred modality in Italy^75^ during spring 2020, but it is not used as extensively in other geographies, mainly applied for patients with CT/CXR contraindications and predestined to study solid organs unlike the lung. Notably, LUS sensitivity was found higher than CXR for COVID-19 diagnosis^76^, and some even found comparable diagnostic accuracy to CT^77,78^. However, the role of LUS for the COVID-19 pandemic is still actively debated^79,80, 81^ and, regarding AI, with only one publicly available dataset^25^ more research is needed to narrow down the practical role of AI on LUS^25,50,82,83^. Additionally, studies using ML on multiple imaging modalities from the same cohort are certainly needed to shed light on comparative questions between modalities from the perspective of ML. The performance of AI-assisted radiologists in detecting COVID-19 might or might not confirm the current radiologic findings, for example that CXR is less sensitive than CT^84^ and LUS (when compared to RT-PCR^85^ or CT^86^) or that B-lines are the most reliable pathological pattern across CT, CXR and LUS^87^. From the AI perspective, LUS is presumably the modality with the highest improvement potential in medical image analysis in the near future. Ultimately, AI technology focusing on plain CXR/LUS data may enable wider leverage in developing countries with limited medical resources

#### 4.1.4 ML interpretability

The combined lack of robustness and interpretability poses steep challenges for the adoption of AI models in clinical practice^88^. Models trained without optimizing for reliability typically make over-confident wrong predictions or under-confident right predictions, especially when extrapolating data. In order to ensure that models make decisions for the right reasons, they must be trained to recognize out-of-distribution samples, and handle distribution shifts. Thereby, allowing models to abstain from making predictions when it is unsure, and deferring such samples to the experts. A human-interpretable access to the model’s decision process is crucial to hone trust in AI, especially in medical applications where reasoning is inductive, sensitive decisions are made and patients expect plausible hypotheses from physicians. In MI, heatmap techniques (like GradCAM ^89^ or guided-backpropagation^90^) and uncertainty estimation of individual predictions (e.g., with MC Dropout^91,92^ or test-time-augmentation ^93^) are the most widely adopted approaches. However, most current interpretability tools focus on generating explanations which highlight patterns learned from the data but do not translate model decisions in human-understandable forms. Counterfactual reasoning has found its way into ML explainability^94^, opened doors toward contrastive explanations (by ascribing how changing the input would affect predictions) and can readily be combined with uncertainty quantification principles to build models integrating reliability into the optimization process^95^. This will enable model introspection and facilitate human-in-the-loop analysis while also considering the performance distribution among human evaluators.

#### 4.1.5 Collaboration between AI and clinical communities

A standard healthcare AI project workflow involves defining a use-case, curating data and annotations, identifying problem constraints, choosing relevant metrics, designing and building the AI system, and lastly evaluating the model performance (see Figure 7 top). However, any problem involves many stakeholders: patients, ethics committee, regulatory bodies, hospital administrators, clinicians and AI experts^97^. In general, data-driven constraints identified by the AI experts tend to transform the clinical task into an evolved task. This in combination with the disconnect of other parties (e.g., clinicians, patients) in the build lifecycle causes potential gaps in the overall outcomes of the collaboration. Awareness and understanding of the difference in needs, motivations, and solution interpretations across agents is imperative. For example, for clinicians, generation of data and metadata are cumbersome, time demanding and tedious. What drives and motivates clinicians are improved clinical workflows and the knowledge and better understanding the analysis can bring, so that they can provide improved patient care. Moreover, AI models may hide inherent risks such as the codification of biases, the weak accountability, and the bare transparency of their decision-making process. Therefore, the way AI models are evaluated can have multiple implications on their applicability, generalization and translation to clinical practice^96,97^. To this end, both the definition of the task to be implemented and evaluated, but also the types of metrics to be leveraged to evaluate the results’ outcomes can be different across collaborators, and hence must be collectively defined. We illustrate such an improved workflow that incorporates other stakeholders in the build process, robust metrics, and iterative useability studies in Figure 7 (bottom). We believe that such a workflow could critically improve the quality of collaboration between AI and clinicians.

**Fig. 7:**
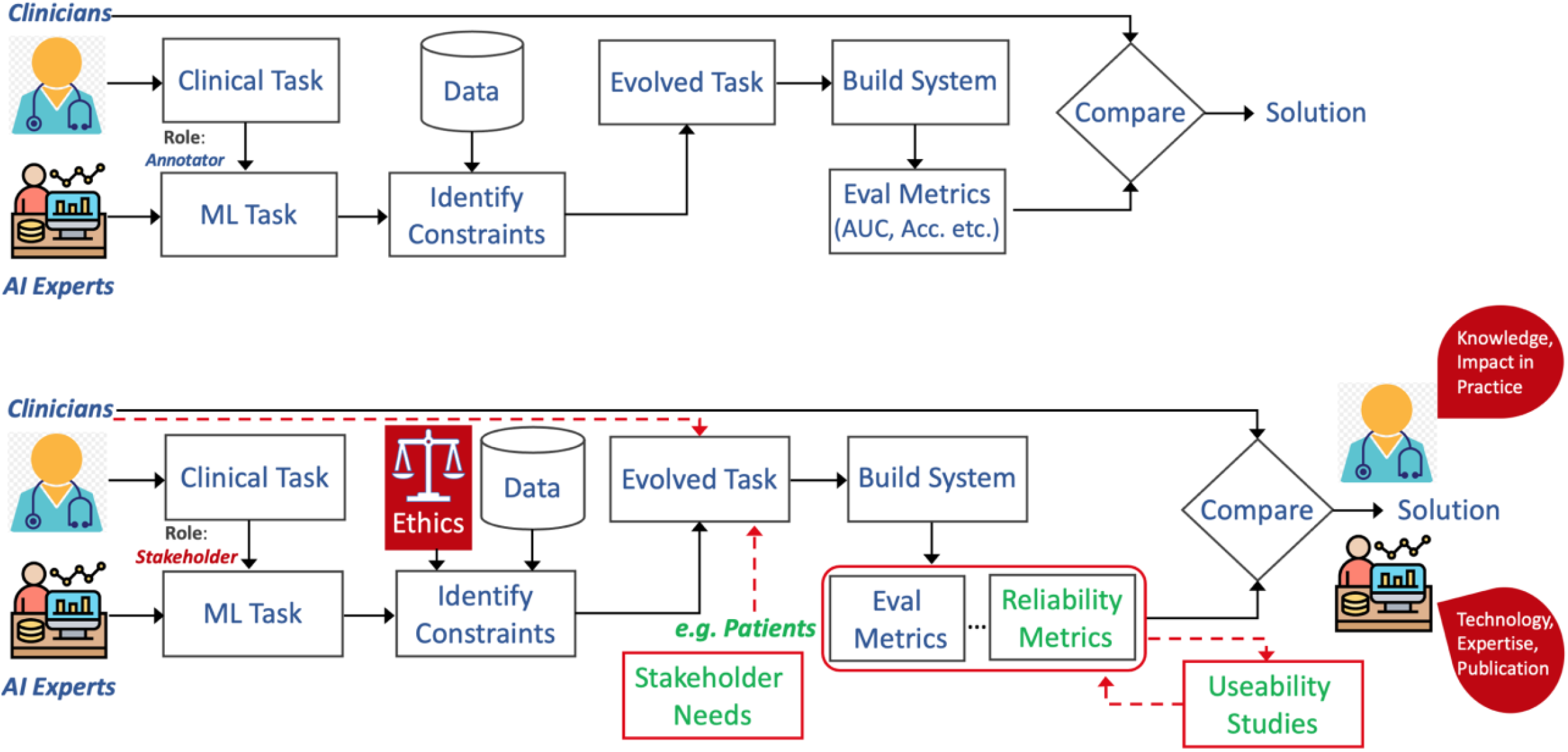
Workflow of collaboration between AI and Clinical experts. (Top) Typical process of developing healthcare AI technology including task definition, data curation, building ML systems, and human-in-the-loop evaluation. (Bottom) Our proposed workflow, highlighting key components that need to be incorporated into the process to improve collaboration between AI and clinical experts. Note the disparity in value interpretation of the developed solutions by the two communities.

To enable agile and transparent development with continuous feedback and evaluation loops, new conducive environments are necessary. A collaboration environment that enables sharing of data, code and results, but also immediate feedback and discussion platforms across collaborators is essential. Communities of discovery such as the digital mammography DREAM challenge^98^ that bring together experts across domains under a unified cloud-based platform can enable data privacy and compliance through distributed and federated learning. Data and code-sharing through open source and open access initiatives, and comprehensive, multidisciplinary validation could pave the way towards closing the gap between technology development and translation to clinical practice.

To summarize, the **challenges** toward improved collaboration include (i) Aligning goals of diverse stakeholders (clinicians, AI experts, patients, funding and regulatory agencies etc.) and (ii) Mapping a medical need into a well-defined task with a measurable and applicable outcome. Possible **solutions** include (i) Inclusive execution and transparency (e.g., keep clinicians and/or patients involved throughout the build process, (ii) Robust evaluation of systems (e.g., going beyond accuracy metrics to incorporate reliability metrics), and (iii) Create common work environments.

Despite the scientometric research which revealed that during COVID-19, global research investments and publication efforts have grown dramatically ^99^, research team sizes, number of involved countries and ratio of international collaborations shrank^100^, we hope to encourage more international collaborations between the AI community and medical experts. This could lead to more mature and conducive technologies and potentially assist clinicians and radiologists in addressing pressing clinical decision support needs during the pandemic.

## Data Availability

No clinical data was used.

## Abbreviations

ACR: (American College of Radiology)
AI: (Artificial Intelligence)
CT: (Computed Tomography)
CXR: (Chest Radiographs)
DL: (Deep Learning)
US: (Ultrasound)
LUS: (Lung Ultrasound)
MI: (Medical Imaging)
PRISMA: (Preferred Reporting Items for Systematic Reviews and Meta-Analyses)
RT-PCR: (Reverse Transcriptase Polymerase Chain reaction)

## Data and code availability

The source code used for the publication keyword search is available via https://pypi.org/project/paperscraper/. A spreadsheet with the detailed results of the publication meta-analysis is enclosed as Supplementary Material (online only).

## Footnotes

* https://pypi.org/project/paperscraper/

