## Appendix for "On the Role of Artificial Intelligence in Medical Imaging of COVID-19"

ScienceDirect

### 1. Appendices

#### Appendix A. Details on meta-review

**Workflow of manual review.**
Fig. 1A (main manuscript) shows how the set of 465 papers for manual review was created. First, a search of the keywords "AI + Medical imaging + COVID-19" revealed 827 matches across the 4 servers *PubMed, arXiv, biorXiv and medRxiv*. Those papers were not specific to lung imaging and thus, a second set of keyword searches were made using "AI + Lung + Imaging + COVID-19 + Modality" where Modality was from {CT, X-Ray, Ultrasound}. This revealed 629 publications (visualized in Fig. 2B). Duplicates were removed and subsequently, the titles were scanned manually and papers that only touched peripherally on AI (or one of the other aspects) were removed. This led to a set of 524 papers. From the 524 manually reviewed papers, 61 were excluded during in-depth analysis since they did not involve any work on AI, leading to the final set of 463 papers.

The manual review was split across 7 authors of this paper (AC, DB, DR, EK, JB, MG, VM) and every paper was evaluated by: primary location of authors, primary location of COVID-19 data, imaging modality, task performed, data origin (external or internal) and maturity assessment.

**Software for keyword search.**
The number of publications per keyword were fetched via a Python package that can be used to reproduce the figures and is publicly available at: https://pypi.org/project/paperscraper/. The queries to the APIs of *PubMed*, *arXiv*, *bioRxiv* and *medRxiv* were made using synonyms or each keyword where a paper was considered a match when title or abstract contained at least one of the synonyms for each keyword (see Table A1). The reference date for all calculations was 31.12.2020.

**Table A1 – List of considered synonyms per search keyword**

| Keyword | Synonyms |
|---|---|
| **COVID-19** | **SARS-CoV-2, corona** |
| **Imaging** | **Image, screen, screening, scan** |
| **Medical imaging** | **Medical image** |
| **AI** | **Artificial intelligence, deep learning, machine learning, neural network, computer vision** |
| **Lung** | **Chest, pulmonary** |
| **Breast** | **Mammography** |
| **CT** | **Computed tomography** |
| **X-Ray** | **XRay, CXR, radiography** |
| **Ultrasound** | **Sonography, LUS** |

**Distributions of modality, task and maturity**
Figure A1 provides details from the meta-review and quantifies how task and maturity are distributed by modality. For example, 87% of all 230 CXR works performed diagnosis whereas this was only the case for 58% of CT papers, where a richer set of papers was found (~15% of works on segmentation and severity assessment, whereas this was 3-4% for CXR). Projects using X-Ray for diagnosis were on average of much lower quality (82% low quality) compared to CT (60%).

Peer review under responsibility of xxxxx.

ELSEVIER Hosting by Elsevier
xxxx-xxxx/$ – see front matter © 2020 xxxxxxxx. Hosting by Elsevier B.V. All rights reserved.

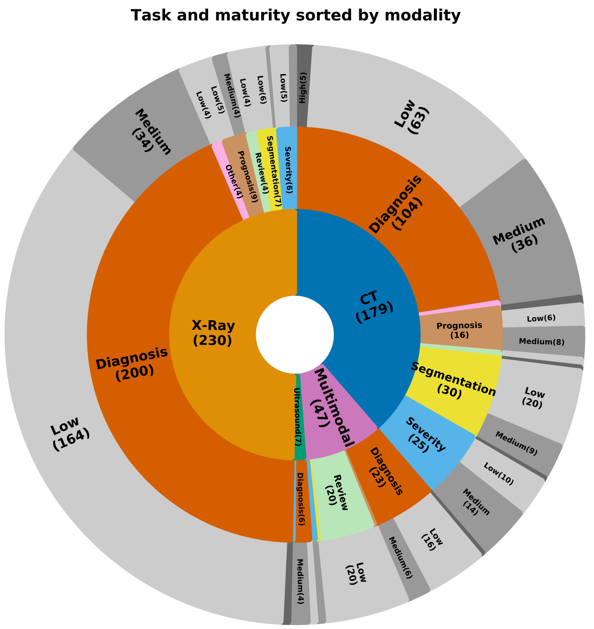


Fig. A1. Sunburst plot on the quality of AI papers distributed by task and modality. The root is in the center and layers are added hierarchically towards the periphery. Hence, the plot should be read inside-out. E.g., From the 179 papers on CT imaging, 104 were dedicated to diagnosis with 63 having low, 36 middle and 5 high maturity. Labels are omitted for fields with less than 5 publications

### Appendix B. Imaging market sizes

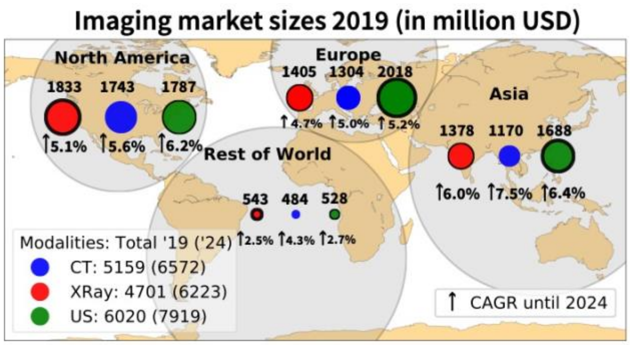


Fig. A2: Regional sizes of medical imaging markets. Overall, US has the largest market across all modalities and is especially dominant in Europe. CT instead is dominant in North America, but CAGR for US are higher and a turn-around is predicted until 2024. The circle radius is proportional to the market size, but for visual clarity, thickness of black borders resembles the regional rank per modality. Figure was created with data from: https://www.marketsandmarkets.com/, Diagnostic Imaging Market - Forecast To 2024.
